# SARS-CoV-2 environmental contamination from hospitalised COVID-19 patients receiving aerosol generating procedures

**DOI:** 10.1101/2021.07.04.21259945

**Authors:** RL Winslow, J Zhou, EF Windle, I Nur, R Lall, C Ji, JE Millar, P Dark, J Naisbitt, A Simonds, J Dunning, W Barclay, K Baillie, GD Perkins, MG Semple, DF McAuley, CA Green

## Abstract

**Background:** Continuous positive airways pressure (CPAP) and high-flow nasal oxygen (HFNO) are considered ‘aerosol-generating procedures’ (AGPs) in the treatment of COVID-19. We aimed to measure air and surface environmental contamination of SARS-CoV-2 virus when CPAP and HFNO were used, compared with supplemental oxygen, to investigate the potential risks of viral transmission to healthcare workers and patients.

**Methods:** 30 hospitalised patients with COVID-19 requiring supplemental oxygen, with a fraction of inspired oxygen ≥0.4 to maintain oxygen saturations ≥94%, were prospectively enrolled into an observational environmental sampling study. Participants received either supplemental oxygen, CPAP or HFNO (n=10 in each group). A nasopharyngeal swab, three air and three surface samples were collected from each participant and the clinical environment. RT qPCR analyses were performed for viral and human RNA, and positive/suspected-positive samples were cultured for the presence of biologically viable virus.

**Results:** Overall 21/30 (70%) of participants tested positive for SARS-CoV-2 RNA in the nasopharynx. In contrast, only 4/90 (4%) and 6/90 (7%) of all air and surface samples tested positive (positive for E and ORF1a) for viral RNA respectively, although there were an additional 10 suspected-positive samples in both air and surfaces samples (positive for E or ORF1a). CPAP/HFNO use or coughing was not associated with significantly more environmental contamination. Only one nasopharyngeal sample was culture positive.

**Conclusions:** The use of CPAP and HFNO to treat moderate/severe COVID-19 was not associated with significantly higher levels of air or surface viral contamination in the immediate care environment.

## INTRODUCTION

Severe acute respiratory syndrome coronavirus 2 (SARS-CoV-2) is a novel betacoronavirus that has led to the global pandemic of Coronavirus disease 2019 (COVID-19), as declared by the World Health Organisation on 11^th^ March 2020. Transmission is by close contact, droplets (>5-10µm diameter) that deposit closer to their source, and airborne inhalation of aerosols (<5µm diameter) that suspend in the air for longer, travel further and have the potential to reach the alveolar region of the lung. Airborne transmission has historically been associated with the use of aerosol generating procedures (AGPs)^1 2^.

UK data from 2020 estimated that 17% of all emergency COVID-19 admissions required respiratory support in high-dependency or intensive-care (ICU) settings, which included the use of non-invasive respiratory support and mechanical ventilation for moderate/severe cases (16% and 10% of all admissions respectively)^3^. Types of non-invasive respiratory support commonly include the use of continuous positive airway pressure (CPAP) and high-flow nasal oxygen (HFNO) devices which have been associated with reductions in mortality and progression to intubation for hypoxemic respiratory failure in some studies^4,5^. Their effectiveness in the treatment of COVID-19 is currently under evaluation in randomised controlled trials. Both are widely designated as AGPs and necessitate additional airborne precautions including cohorting of patients and the use of FFP3 masks for healthcare workers (HCWs) to mitigate the risk of aerosol transmission^6 7^. However this is based on weak evidence from the SARS-CoV-1 outbreak and may delay or restrict patient access to the use of these therapies ^8^. Nosocomial transmission from earlier coronavirus outbreaks (SARS-CoV-1 and MERS-CoV) were reported as up to 80% and 40% for patients and HCWs respectively^9^ and recent studies suggest that HCWs represent a population with a substantial burden from COVID-19, particularly in non-ICU settings where airborne precautions are less frequently used^10 11^.

SARS-CoV-2 environmental contamination has been widely found in multiple studies, however very few have specifically evaluated the impact of CPAP and/or HFNO, or have found biologically viable virus that proves a transmission risk to HCWs^12-20^. Other studies in this field include aerosol generation studies that have mainly used patient simulators or healthy volunteers^21-24^. Here we report our observations from sampling the clinical environment of COVID-19 patients undergoing CPAP and HFNO, compared to the use of supplemental oxygen, to better understand the risks of airborne and fomite SARS-CoV-2 contamination and exposure to HCWs.

## MATERIALS & METHODS

### Study design, participants and setting

This study was a prospective observational study of environmental viral contamination from hospital admissions with COVID-19 as part of the International Severe Acute Respiratory and emerging Infections Consortium (ISARIC) WHO Clinical Characterisation Protocol UK (CCP-UK, www.isaric4c.net). It was performed across three UK hospitals at University Hospitals Birmingham NHS Foundation Trust and study participants were NHS patients co-enrolled (or who were eligible to be co-enrolled) into ISARIC WHO CCP-UK and the RECOVERY-Respiratory Support trial^25^. Participant inclusion criteria included having suspected or confirmed SARS-CoV-2 infection with hypoxaemia (defined as requiring supplemental oxygen with a fraction of inspired oxygen ≥0.4 to maintain oxygen saturations ≥94%) and suitable for CPAP or HFNO. Participants were enrolled into one of three groups; CPAP, HFNO or supplemental oxygen, within 5-days of commencing treatment. Recruitment was opportunistic and written informed consent was obtained before any study procedures were undertaken. The machines used to deliver CPAP were either a Philips Respironics Trilogy, V60 using ResMed AcuCare masks with Heat Moisture Exchange filter (HME), or the University College London (UCL) Ventura system with viral filters, and all were capable of flow rates from 15-60L/min. HFNO was delivered by a Fisher and Paykel Airvo2 system using Optiflow nasal cannulae (OPT944) with a typical flow rate between 50-60L/minute. Participants received supplemental oxygen via a Venturi facemask with a maximal flow of 15L/min. The flow rate, inspired FiO_2_ and positive end expiratory pressures (PEEP) were set according to clinical need.

### Data and sample collection

Environmental samples were taken from the care setting of each participant, which varied according to clinical and operational needs. Basic demographic and clinical data were collected with samples in a single visit that lasted up to 60-minutes. Room temperature, humidity and carbon dioxide levels were recorded using a Therm M2000C Air Quality Monitor. Nasopharyngeal samples were collected using a mid-turbinate flocked swab in accordance with standard operating procedures and stored in viral transport medium (VTM). Air samples were collected using a Coriolis micro (μ) air sampler (Bertin Technologies, France) that uses liquid cyclonic technology able to collect particles from 0.5μm in diameter^16^. The device inlet was aligned to the mouth of the participant at a distance of 50cms, and sampled the air on three occasions, each for 10-minutes at a flow rate of 300L/min (total 9m^3^ air). The first air sample was collected with the participant at rest with supplemental oxygen only. Where the participant was unable to tolerate removal of CPAP/HFNO for the first sample, this was collected on CPAP/HFNO in order to keep the sampling period consistent for all participants. The next air sample was with CPAP/HFNO in place for a minimum of 5-minutes (or supplemental oxygen) and the third air sample involved the addition of voluntary coughing every 2-minutes. All surface samples were taken from within 2m of the participant and used sterile flocked swabs (Coplan, US) pre-moistened with VTM to swab 25cm^2^ from the floor, the bed table and a high-object (above participant head height such as a light fitting), in accordance with WHO sampling guidance^26^. All swabs were placed into 1mLs VTM. All samples were stored on ice for less than 2-hours before being stored at -80 °C and later transported in accordance with UN3373 using chilled biotherm containers that maintained storage temperature at 4-6 °C for laboratory analysis at Imperial College London.

### Detection and quantification of human and SARS-CoV-2 viral RNA by real-time polymerase chain reaction and viral cultures

Laboratory analyses were performed blinded to study group. Viral RNA detection and quantification was performed using quantitative real-time reverse transcription polymerase chain reaction (RT-qPCR), as described elsewhere^16^. In summary, samples were extracted from 200µL of the VTM medium using the QIAsymphony SP (Qiagen, Germany) instrument according to the manufacturer’s instructions and SARS-CoV-2 viral RNA was detected using AgPath-ID One-Step RT-PCR Reagents (Life Technologies) with specific primers and probes targeting the envelope (E)^27^ and ORF1a genes. A standard curve with six serial dilutions of 1×10^5^ – 1 x 10^0^ copies/ul E gene was included in each run of the RT-qPCR. A sample was defined as positive for viral RNA if both E and ORF1a RT-qPCR assays gave cycle time (Ct) values <45. A Ct value <45 for only one of these viral gene targets was considered a suspected-positive result. A one-step RT-qPCR assay targeting human RNaseP was used to indicate human biological material in nasopharyngeal and surface swabs^28^. Human biological material in air samples was quantified by a one-step RT-qPCR assay targeting human 18s rRNA (18s rRNA_Forward 5’-GGTAACCCGTTGAACCCCAT-3’, 18s rRNA_Reverse 5’-CAACGCAAGCTTATGACCCG-3’, 18s rRNA_Probe 5’-FAM-GTGATGGGGATCGGGGATTG-BHQ1-3’). A sample was defined as positive for human RNA if the Ct value was <45. Vero E6 (African Green monkey kidney) cells were used to culture virus from any positive/suspected positive viral RNA sample. Vero were maintained in DMEM supplemented with heat-inactivated foetal bovine serum (10%) and Penicillin/Streptomycin (10, 000 IU/mL &10, 000 µg/mL). For virus isolation, 200 µL of samples were added to 24 well plates. On day 0 and after 5-7 days the cell supernatants were collected, and RT-qPCR used to detect SARS-CoV-2 RNA as described above. Samples with at least one log increase in copy numbers for the E gene (reduced Ct values relative to the original samples) after 5-7 days propagation in cells compared with the starting value were considered positive by viral culture^29^.

### Statistical analyses

This was an exploratory study intended to be descriptive in nature, no formalised sample size was calculated, and a sample size of 30 was chosen to fulfil the criteria of the central limit theorem which allows for stability in the estimates (mean and standard deviation) of the outcomes^30^. Analysis of variance (ANOVA/Kruskal-Wallis, as appropriate) were used to provide an overall comparison of the three groups, and significant variations were further explored by pairwise comparisons (unpaired t-tests against the supplemental oxygen group). All statistical tests were two-tailed and a p-value<0.05 was considered statistically significant. The statistical models were fitted in SAS and Prism 7 (GraphPad Inc, USA). Statistically significant differences should be interpreted with caution as the study was not powered to detect differences in the treatment arms.

## RESULTS

32 eligible patients were invited to take part and two declined to participate. Samples from the 30 enrolled participants were collected between 11/12/2020 and 19/02/2021, when the dominant variant was likely to have been B.1.1.7. The study population demographics, clinical characterisation of COVID-19 disease, and environmental conditions of the care provided to them are presented in Table 1. All participants required oxygen support on admission and commenced dexamethasone the same day. Participant demographics were comparable across the study groups. Participants from the HFNO group were sampled significantly later in their illness compared with those receiving supplemental oxygen (mean 16 days, 95%CI 13-19, vs mean 9 days, 95%CI 5-13, from symptom onset respectively) and participants receiving supplemental oxygen were sampled significantly earlier into their hospital stay (median 1-day, IQR 0-3, compared with CPAP median of 4.5 days, IQR 2-6, and HFNO median of 3-days, IQR 2-6) (sFig. 1). Similar proportions of patients in each study group were cared for in cohorted areas or side room settings. Participants receiving CPAP/HFNO were more commonly accommodated in negative-pressure rooms. Compared with patients receiving supplemental oxygen, the room air recordings measured significantly lower temperatures for HFNO, with lower CO_2_ content and humidity for CPAP (sFig. 2).

**Table 1.** The baseline clinical characteristics of study participants and the environment of care provision. A total of 30 participants with moderate/severe COVID-19 were enrolled into the study. Paired t-tests were post-hoc analysis of differences between SOC and CPAP/HFNO study groups only. SOC, supplemental oxygen care. CPAP, continuous positive airway pressure. HFNO, high-flow nasal oxygen. FiO_2_, fraction of inspired oxygen. SpO_2_, oxygen saturation. CI, confidence interval. IQR, interquartile range. n/a, not applicable. ns, not significant.

|  | All | SOC | CPAP | HFNO | Statistically significant differences |
| --- | --- | --- | --- | --- | --- |
| **Number of participants** | 30 | 10 | 10 | 10 | - |
| **n Male gender** | 17 | 6 | 5 | 6 | - |
| **Mean age** [95% CI mean]<br>(min-max) | 56 [52-60]<br>(35-75) | 54 [47-61]<br>(35-74) | 60 [52-68]<br>(44-75) | 53 [45-61]<br>(39-68) | p=ns (ANOVA) |
| **Ethnicity** |  |  |  |  |  |
| n Asian – Pakistani | 10 | 2 | 6 | 2 | - |
| n White -British | 8 | 4 | 0 | 4 | - |
| n Not given | 4 | 1 | 3 | 0 | - |
| n Asian - Indian | 3 | 0 | 0 | 3 | - |
| n Asian - Other | 2 | 1 | 0 | 1 | - |
| n White - Other | 1 | 1 | 0 | 0 | - |
| n Caribbean | 1 | 1 | 0 | 0 | - |
| n Mixed – White and Caribbean | 1 | 0 | 1 | 0 | - |
| **Mean number of days of illness at time of hospital admission**<br>[95% CI mean] (min-max) | 9 [8-11]<br>(0-17) | 8 [5-11]<br>(2-15) | 8 [6-11]<br>(3-12) | 11 [8-14]<br>(0-15) | p=ns (ANOVA) |
| **Mean number of days of illness at time of sampling**<br>[95% CI mean] (min-max) | 12 [10-14]<br>(3-25) | 9 [5-13]<br>(3-18) | 13 [9-16]<br>(6-24) | 16 [13-19]<br>(11-25) | p=0.02 (ANOVA)<br>SOC vs CPAP p=ns (unpaired t-test)<br>SOC vs HFNO p<0.01 (unpaired t-test) |
| **Median number of days in hospital at the time of sampling**<br>[IQR] (min-max) | 2 [1-5]<br>(0-14) | 1 [0-2]<br>(0-3) | 4.5 [2-6]<br>(1-9) | 3 [2-6]<br>(2-14) | p<0.01 (Kruskal-Wallis)<br>SOC vs CPAP p<0.01 (Mann-Whitney)<br>SOC vs HFNO p<0.01 (Mann-Whitney) |
| **Mean number of days CPAP/HFNO at time of sampling**<br>[95% CI mean] (min-max) | n/a | n/a | 2.4 [1.5-3.3]<br>(1-4) | 1.8 [1.0-2.6]<br>(0-3) | - |
| **Median FiO<sub>2</sub> at time of sampling**<br>[IQR] (min-max) | 56 [40-73]<br>(35-98) | 59 [40-65]<br>(40-98) | 48 [40-62]<br>(40-80) | 63 [40-91]<br>(35-98) | p=ns (Kruskal-Wallis) |
| **Mean SpO<sub>2</sub> at time of sampling**<br>[95% CI mean] (min-max) | 94 [93-95]<br>(92-99) | 95 [93-96]<br>(92-99) | 94 [93-95]<br>(92-96) | 94 [93-96]<br>(92-98) | p=ns (ANOVA) |
| **Room type** |  |  |  |  |  |
| Open bay/cohort area | 12 | 4 | 4 | 4 | - |
| Side room – ambient pressure | 8 | 5 | 0 | 3 | - |
| Side room – negative pressure | 7 | 0 | 6 | 1 | - |
| Side room – natural airflow | 3 | 1 | 0 | 2 | - |
| **Estimated air changes per hour (ACH)** |  |  |  |  |  |
| 10 ACH | 15 | 6 | 6 | 6 | - |
| 4 to 6 ACH | 10 | 4 | 4 | 2 | - |
| 4 ACH | 2 | 0 | 0 | 2 | - |
| **Mean room air temperature (°C) at time of sampling**<br>[95% CI mean] (min-max) | 21.9 [21-23]<br>(18.0-25.0) | 23.2 [22-24]<br>(20.0-25.0) | 21.9 [21-23]<br>(19.0-24.0) | 20.7 [19-22]<br>(18.0-23.0) | p=0.01 (ANOVA)<br>SOC vs CPAP p=ns (unpaired t-test)<br>SOC vs HFNO p<0.01 (unpaired t-test) |
| **Median room air CO<sub>2</sub> content (ppm) at time of sampling**<br>[IQR] (min-max) | 574.5<br>[500-808]<br>(419-1548) | 672.5<br>[530-774]<br>(459-1548) | 502.0<br>[448-582]<br>(419-618) | 915.0<br>[459-1303]<br>(506-1460) | p=0.01 (Kruskal-Wallis)<br>SOC vs CPAP p=0.02 (Mann-Whitney)<br>SOC vs HFNO p=ns (Mann-Whitney) |
| **Mean room air humidity (%) at time of sampling**<br>[95% CI mean] (min-max) | 37.6 [34-41]<br>(22.0-58.0) | 37.5 [32-43]<br>(23.0-41.0) | 30.4 [26-35]<br>(23.0-41.0) | 44.8 [37-53]<br>(26.0-58.0) | p<0.01 (ANOVA)<br>SOC vs CPAP p=0.03 (unpaired t-test)<br>SOC vs HFNO p=ns (unpaired t-test) |
| **n Receiving humidified oxygen** | 15 | 6 | 2 | 7 | - |
| **n CPAP full facemask (un-vented)** | n/a | n/a | 8 | n/a | - |
| **n CPAP partial facemask (vented)** | n/a | n/a | 2 | n/a | - |

### Participants had detectable viral RNA in the nasopharynx at the time of environmental sampling

Overall 21/30 (70%) of participants tested positive for SARS-CoV-2 RNA in the nasopharynx at the time of environmental sampling (Table 2). An additional participant was a suspected-positive case and all study participants tested positive on PCR testing either in the community or on admission to hospital (data not shown). For positive samples, the mean Ct value was 29.2 (95%CI 27-32) and were comparable across different study groups with high correlation between the Ct value for each gene (r^2^=0.95). There were no correlations between the Ct values of any viral genes and the duration of illness/hospital stay (sFig. S3).

**Table 2.** The frequencies of SARS-CoV-2 RNA positive, suspected-positive and negative samples. A Ct value <45 for both the SARS-CoV-2 E gene and ORF1a gene was considered a positive result. A suspected positive result was recorded when only E or ORF1a Ct values were <45. A negative result was recorded when both E and ORF1a Ct values were ≥ 45. Nasopharyngeal samples were collected according to local standard operating procedures and air samples and surfaces samples were collected per participant in accordance with the clinical study plan. There were no statistically significant differences in the Ct values of viral RNA in nasopharyngeal samples between study groups (p=ns, two-way ANOVA), and no statistically significant differences in the proportion of negative samples in each air and surface sample across the study groups (p=ns, Fisher’s exact). Alternative statistical tables are available in supplementary material. SOC, supplemental oxygen care. CPAP, continuous positive airway pressure. HFNO, high-flow nasal oxygen. CI, confidence interval.

|  | All | SOC | CPAP | HFNO |
| --- | --- | --- | --- | --- |
| **Number of participants** | 30 | 10 | 10 | 10 |
| **Nasopharyngeal samples** |  |  |  |  |
| Number positive/suspected-positive/negative (overall % positive or suspected-positive)<br>Mean Ct value [95% CI] for lowest Ct value for E or ORF1a only | 21/1/8 (73%)<br>29.2 [27-32] | 8/1/1 (90%)<br>29.8 [26-34] | 8/0/2 (80%)<br>31.2 [27-35] | 5/0/5 (50%)<br>24.9 [18-32] |
| **Overall for air samples** |  |  |  |  |
| Number positive/suspected-positive/negative (overall % positive or suspected-positive) | 4/10/76 (16%) | 1/4/25 (17%) | 0/2/28 (7%) | 3/4/23 (23%) |
| **Air samples collected with participant breathing normally (SOC or CPAP/HFNO off)** |  |  |  |  |
| Number positive/suspected-positive/negative (overall % positive or suspected-positive)<br>Mean Ct value [95% CI] for lowest Ct value for E or ORF1a only | 2/4/24 (20%)<br>38.2 [35-41] | 1/2/7 (30%)<br>39.7 [32-48] | 0/1/9 (10%)<br>37.3 [-] | 1/1/8 (20%)<br>36.3 [7-66] |
| **Air samples collected with participant breathing normally (SOC or CPAP/HFNO on)** |  |  |  |  |
| Number positive/suspected-positive/negative (overall % positive or suspected-positive)<br>Mean Ct value [95% CI] for lowest Ct value for E or ORF1a only | 1/3/26 (13%)<br>39.0 [34-44] | 0/1/9 (10%)<br>37.4 [-] | 0/0/10 (0%)<br>- [-] | 1/2/7 (30%)<br>39.6 [31-48] |
| **Air samples collected with participant coughing every 2min (SOC or CPAP/HFNO on)** |  |  |  |  |
| Number positive/suspected-positive/negative (overall % positive or suspected-positive)<br>Mean Ct value [95% CI] for lowest Ct value for E or ORF1a only | 1/3/26 (13%)<br>38.6 [35-42] | 0/1/9 (10%)<br>39.9 [-] | 0/1/9 (10%)<br>39.9 [-] | 1/1/8 (20%)<br>37.5 [13-63] |
| **Overall for surface samples** |  |  |  |  |
| Number positive/suspected-positive/negative (overall % positive or suspected-positive) | 6/10/74 (18%) | 1/4/25 (17%) | 3/3/24 (20%) | 2/3/25 (17%) |
| **Floor surfaces** |  |  |  |  |
| Number positive/suspected-positive/negative (overall % positive or suspected-positive)<br>Mean Ct value [95% CI] for lowest Ct value for E or ORF1a only | 5/4/21 (30%)<br>37.3 [36-48] | 1/1/8 (20%)<br>35.8 [18-54] | 3/1/6 (40%)<br>36.8 [36-38] | 1/2/7 (30%)<br>38.9 [37-41] |
| **Table surfaces** |  |  |  |  |
| Number positive/suspected-positive/negative (overall % positive or suspected-positive)<br>Mean Ct value [95% CI] for lowest Ct value for E or ORF1a only | 0/3/27 (10%)<br>39.0 [36-42] | 0/2/8 (20%)<br>38.5 [30-47] | 0/0/10 (0%)<br>- [-] | 0/1/9 (10%)<br>40.0 [-] |
| **High-object surfaces** |  |  |  |  |
| Number positive/suspected-positive/negative (overall % positive or suspected-positive)<br>Mean Ct value [95% CI] for lowest Ct value for E or ORF1a only | 1/3/26 (13%)<br>37.8 [35-41] | 0/1/9 (10%)<br>39.4 [-] | 0/2/8 (20%)<br>38.4 [32-45] | 1/0/9 (10%)<br>34.8 [-] |

### Low-levels of viral RNA in air samples, regardless of whether CPAP or HFNO was in use or if the participant was coughing

Overall 9/30 (30%) of participants had at least one positive or suspected-positive result from one or more of the three air samples collected (Fig. 1, sFig. S4 and sFig. S5). There were only 4/90 (4%) positive air samples, with an additional 10 suspected-positive. Furthermore, the Ct values for positive and suspected-positive air samples were substantially higher than paired samples in the nasopharynx, indicating minimal viral RNA in the air. The distribution of these positive and suspected-positive air samples did not indicate a relationship with the use of CPAP or coughing, but 7/14 (50%) of the positive and suspected-positive air samples were from the HFNO group despite only half of these participants testing positive for viral RNA on nasopharyngeal samples, although this was not statistically significant (Table 2, Fig. 1). Human 18s RNA was detectable in 85/90 (94%) of air samples. Again, the use of CPAP/HFNO and/or coughing did not appear to alter the quantity of human RNA. Post-hoc analyses explored potential differences between the nine participants who had tested positive or suspected-positive for viral RNA in one or more of the air samples, compared with the other 21 participants with negative air samples. Irrespective of the use of CPAP/HFNO at rest or on coughing, we found no significant differences with the environmental variables, days unwell at time of sampling, or nasopharyngeal Ct values between those who did and did not have viral RNA in air samples.

**Fig. 1.**
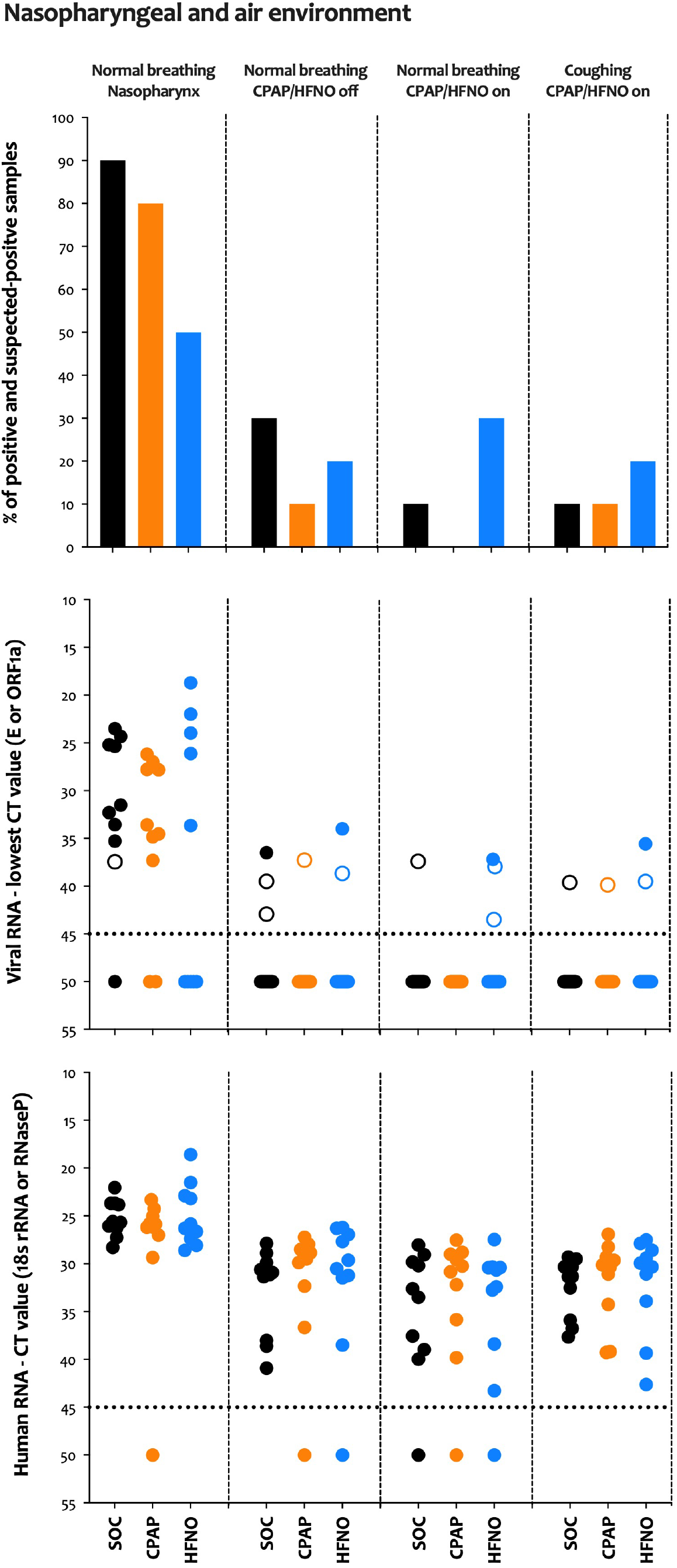
Viral and human RNA from nasopharyngeal and air samples. A total of three air samples were collected per participant. The first was at rest with the patient receiving supplementary oxygen via a face-mask if able to tolerate a pause in CPAP/HFNO treatment for volunteers in these groups. The second sample was at rest with the CPAP/HFNO device on (if applicable). The third sample included voluntary coughing every 2-minutes with the CPAP/HFNO device on (if applicable). (**Top**) The proportion of samples that tested positive or suspected-positive for viral RNA. (**Middle**) Ct values for viral RNA. The dotted line signifies the detection threshold of 45, with Ct values ≥ 45 were considered negative and were arbitrarily assigned a value of 50. Coloured circles show positive results (Ct value <45 in both E and ORF1a genes), whereas empty circles show suspected-positive results (a Ct value <45 in one of the two genes only) (**Bottom**) Ct values for human RNaseP in nasopharyngeal samples and human 18s rRNA in air samples. The dotted line signifies the detection threshold of 45, with Ct values ≥ 45 were considered negative and arbitrarily assigned a value of 50. SOC, supplemental oxygen care. CPAP, continuous positive airway pressure. HFNO, high-flow nasal oxygen.

### Clinical surfaces were more contaminated with viral RNA than the air samples

A higher proportion, 14/30 (47%), of participants had at least one positive or suspected-positive sample for viral RNA from one or more of the three surface samples collected (Fig. 2, sFig. S6 and sFig. S7). Only four participants had a positive or suspected-positive sample in both an air and surface sample (two participants receiving supplemental oxygen and one from CPAP and HFNO). In total, 6/90 (7%) of surface swabs were positive for viral RNA; 5/30 (17%) floor samples tested positive (and 4 suspected-positive), no table surface samples tested positive (and 3 suspected-positive) and only one high-object surface sample tested positive (and 3 suspected-positives). As with our air samples, the Ct values for viral genes were greater than those recorded from the nasopharynx and there were no differences with the use of CPAP/HFNO on any surface type. The floor was the most frequently contaminated surface (30%) followed by the high-object surfaces (13%) and tables (10%). Human RNA could be detected in 28/30 (93%) floor samples, 16/30 (53%) table samples and only 10/30 (33%) high-object surface samples. The Ct values for human RNaseP steadily increased from nasopharyngeal samples to floor, table and then the high-object samples. As before, the subset of participants with one or more positive or suspected-positive surface sample for viral RNA (n=14) were compared against participants who had negative surface swabs (n=16). The Ct values for viral RNA did not appear to vary significantly with the number of days unwell or nasopharyngeal Ct values between those who did and did not have viral RNA in surface samples. Lower room humidity was more common with positive surface samples and no significant differences were observed with other environmental measures.

**Fig. 2.**
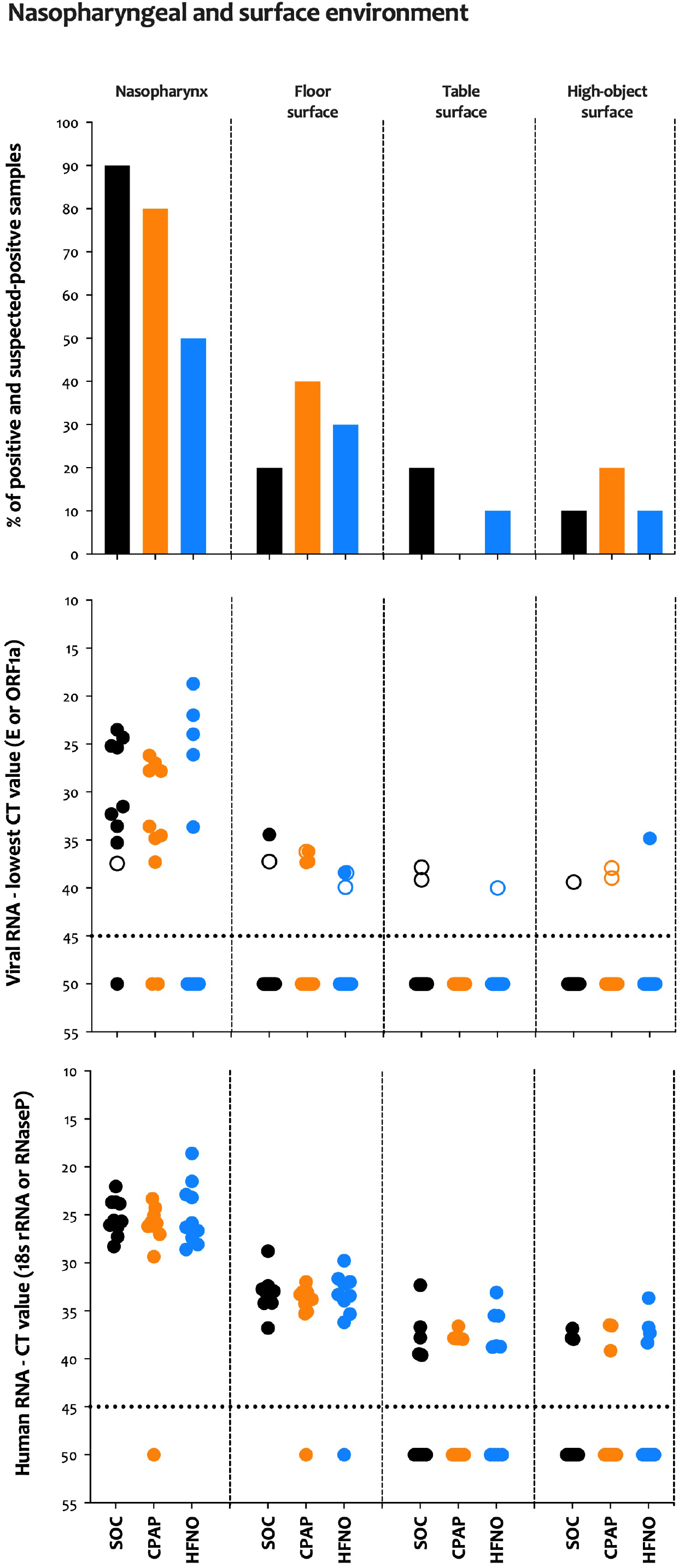
Viral and human RNA from surface samples. A total of three surface samples were collected per participant. The first was from the floor within 2m of the bed, the second sample was from the bedside table at head-height to the participant, and the third sample was from an object above participant head height (for example, a light fitting). (**Top**) The proportion of samples that tested positive or suspected-positive for viral RNA. (**Middle**) Ct values for viral RNA. The dotted line signifies the detection threshold of 45, with Ct values ≥ 45 were considered negative and were arbitrarily assigned a value of 50. Coloured circles show positive results (Ct value <45 in both E and ORF1a genes), whereas empty circles show suspected positive results (a Ct value <45 in one of the two genes only). (**Bottom**) Ct values for human RNaseP in surface samples. The dotted line signifies the detection threshold of 45, with Ct values ≥ 45 considered negative and were arbitrarily assigned a value of 50. SOC, supplemental oxygen care. CPAP, continuous positive airway pressure. HFNO, high-flow nasal oxygen.

### No viable virus could be recovered from any environmental sample that tested positive by PCR

In total, 51/210 (24%) samples were positive or suspected-positive for viral RNA and were cultured. Only one nasopharyngeal sample from a HFNO participant (E gene Ct 21.99) could demonstrate presence of viable (infective) virus and all other samples, including environmental samples, were negative. This individual had two positive air samples that had higher Ct values for viral RNA and were culture negative.

## DISCUSSION

Our sampling study of the immediate environment of patients requiring non-invasive respiratory support for life-threatening COVID-19 disease found that few air and surface samples had measurable viral RNA contamination, irrespective of using CPAP/HFNO and/or coughing. Furthermore, the samples that did detect viral RNA by RT-qPCR, including those from the nasopharynx, failed to demonstrate biological viability in cell-culture except for one nasopharyngeal sample. These data question the infectivity of patients admitted to hospital at this stage of disease, and any additional risks to HCWs/other patients associated with the use of CPAP and HFNO which are considered ‘aerosol-generating’, compared to the use of supplemental oxygen.

Consistent with other environmental sampling studies we found airborne and surface viral RNA contamination, 4% and 7% positive samples respectively, within the vicinity of COVID-19 patients although the degree of contamination is lower than that reported in most other studies^12-20^. This was despite the majority of our participants having detectable viral RNA in the nasopharynx at time of sampling and irrespective of respiratory support type and/or coughing. Importantly few previous studies included patients receiving non-invasive respiratory support, and from those that did there was little or no air contamination around NIV or HFNO^19,16^,^18^. Furthermore, our findings concur with other studies that report surface contamination is not associated with mode of respiratory support including HFNO and/or NIV^12 17^. Consistent with others we found higher rates of floor contamination compared to other surfaces^13 15^. This is unsurprising given the likely cumulative deposition of virus laden droplets from the air combined with potential transference of the virus from footwear. Heterogeneity between clinical setting, study design and methodology limit direct comparisons and is likely to account for the variation in findings between studies.

The lower degree of environmental contamination we found may be related to the stage of disease in our cohort of participants, with one sampling study reporting a decline in environmental contamination after the first week of illness^13^. Participants in our study were on average in their second week of illness when admitted to hospital (mean 9-days) and when sampled (mean 12-days). SARS-CoV-2 viral shedding is at its highest quantity in early infection and the peak of infectivity coincides with symptom onset before a gradual decline to near the detection limit by day 21, albeit with significant individual variability^31-34^. This kinetic is notably different to the related SARS-CoV-1 virus where viral shedding peaks 7-10 days after symptom onset^35 36^, and coincides more with the time when patients are admitted for hospital care. The SARS outbreak was associated with a high incidence of healthcare worker and nosocomial transmission^9^. Although we found no significant relationship between nasopharyngeal viral load and days of illness (or environmental contamination), COVID-19 patients requiring non-invasive respiratory support are more likely to be at a stage of disease when it is plausible that host immunity has begun to establish control of viral shedding and infectivity.

The levels of environmental contamination in our study were not significantly influenced by CPAP/HFNO therapies and/or coughing. These findings broadly reflect data from aerosol generation studies in healthy adult volunteers which report non-invasive positive pressure ventilation (NIPPV) and HFNO did not generate significantly more aerosols (compared to other respiratory activities)^21 24^ or in fact reduced emissions for NIPPV and HFNO^22^ and CPAP^23^. This may be influenced by the semi-closed system of CPAP delivery and PEEP over the nose and mouth simultaneously that limit aerosol/droplet dispersion from respiratory secretions. High-flow nasal cannulae to deliver HFNO leaves the mouth open for potential expulsion of infective secretions. Hamilton et al report HFNO was associated with increased aerosol emission (flow rate and machine dependent), however this was generated by the machine, not the patient, hence unlikely to carry SARS-CoV-2 virus. Moreover, these studies consistently reported the highest aerosol emissions were from coughing, irrespective of respiratory support modality, with at least a 3-fold increase^21-23^. We did not find this signal in our data however these findings indicate that coughing is potentially the most hazardous source of infectious SARS-CoV-2 aerosols to HCWs and not the respiratory support device itself. The extrapolation of data from healthy volunteers may be limited to COVID-19 patients, however one study has shown that the aerosol particle size distribution is similar between the two populations^23^. Collectively, data from these studies and our own findings question whether the airborne mitigation measures are correctly aligned to the highest transmission risk, most likely from coughing and not the form of non-invasive respiratory support used.

Importantly, we found no biologically viable virus in cell culture from any positive or suspected-positive environmental samples except for one nasopharyngeal sample from a HFNO participant (E gene Ct value 21.99). This was a common finding from other environmental sampling studies that attempted culture^12 16,14 18 20^. This may be due to air sampling methods which are known to inactivate viruses and impact upon virus infectivity^37 38^ although all of our surface and nasopharyngeal samples (except one) were also negative on cell culture. Lower Ct values have been correlated with a higher likelihood of successful culture^34 39^ with studies demonstrating viable virus could only be cultured from clinical samples and experimentally contaminated surfaces if the Ct value <24 and <30 respectively ^16 40^. All of our positive/suspected-positive environmental samples had a Ct value >30 and were likely to be below the detection threshold. This indicates that not only was there a poverty of viral RNA in the immediate environment of COVID-19 patients receiving respiratory support therapies, but also there was no detectable viable virus present as an infection risk to HCWs.

Our study has some notable strengths and limitations. Strengths include the ‘real-world’ setting, a standardised sampling strategy, concurrent air and surface sampling, collection of patient data and nasopharyngeal samples to understand the clinical context, and the use of human genetic material as a control. Finally, embedding the evaluation within the RECOVERY-Recovery Support randomised controlled trial helped to minimise selection bias. Limitations include the lack of serial sampling with findings representing a ‘snap shot’ picture, potential cross-contamination by other infected patients in cohorted areas, no particle size fractionation or concentration measurement (hence not able to differentiate between droplets and aerosols), air volume sampled only a small fraction of the total room air, and challenges interpreting the significance of samples with low viral loads. The small group sizes risk the study being underpowered with confounding chance observations and larger studies are needed to develop the evidence-base needed to reliably inform pragmatic infection prevention control measures around the use of CPAP/HFNO.

## Conclusions

We found limited SARS-CoV-2 viral RNA within the immediate environment of hospitalised COVID-19 patients and that this was not significantly influenced by the use of CPAP/HFNO devices or coughing, and importantly no detectable biologically viable virus. This adds to an increasing evidence base that in the context of COVID-19, CPAP and HFNO may not be higher transmission risk procedures that are associated with their ‘aerosol generating’ classification.

## Data Availability

Data will be available to researchers on request subject to sponsor approval.

## SUPPLEMENTARY TABLES

**sTab. 1.** Additional post-hoc analytical results for laboratory data. In this analysis, all Ct values >45 were not included. Test is not applicable when number of observations is less than number of treatment groups or there is no valid observation in at least one arm. Overall difference across treatment arms was assessed using Exact Kruskal Wallis test. The interpretation from using this alternative statistical analysis were not different from those presented in the main paper. SOC, supplemental oxygen care. CPAP, continuous positive airway pressure. HFNO, high-flow nasal oxygen. n/a, not applicable.

| Sample | Gene type |  | Treatment arm |  |  | Total | Exact Kruskal Wallis test |
| --- | --- | --- | --- | --- | --- | --- | --- |
|  |  |  | CPAP | HFNO | SOC |  | P value |
| Nasopharyngeal swabs | E gene | N of positive value | 8 | 5 | 9 | 22 | 0.088 |
|  |  | Mean | 31.1 | 24.9 | 29.8 | 29.2 |  |
|  |  | Standard deviation | 4.4 | 5.6 | 5.3 | 5.4 |  |
|  |  | Median | 30.7 | 24 | 31.5 | 27.8 |  |
|  |  | Interquartile range | 27.4, 34.7 | 22.0, 26.1 | 25.2, 33.6 | 25.2, 33.7 |  |
|  |  | Negative value | 2 | 5 | 1 | 8 |  |
|  | ORF1a gene | N | 8 | 5 | 8 | 21 | 0.089 |
|  |  | Mean | 32.2 | 26 | 30.2 | 29.9 |  |
|  |  | Standard deviation | 4.6 | 5.9 | 4.9 | 5.3 |  |
|  |  | Median | 31.5 | 25 | 30 | 28.3 |  |
|  |  | Interquartile range | 28.1, 35.8 | 22.8, 27.0 | 26.0, 34.3 | 26.1, 35.0 |  |
|  |  | Negative value | 2 | 5 | 2 | 9 |  |
|  | RNaseP gene | N | 9 | 10 | 10 | 29 | 0.855 |
|  |  | Mean | 25.9 | 24.9 | 25.2 | 25.3 |  |
|  |  | Standard deviation | 1.7 | 3.2 | 1.9 | 2.4 |  |
|  |  | Median | 25.9 | 26.1 | 25.6 | 25.8 |  |
|  |  | Interquartile range | 25.0, 26.2 | 22.9, 27.4 | 23.7, 26.2 | 23.7, 26.6 |  |
|  |  | Negative value | 1 | 0 | 0 | 1 |  |
| Air sample (1st) | E gene | N | 1 | 2 | 3 | 6 | 0.133 |
|  |  | Mean | 37.3 | 37.7 | 40.6 | 39.1 |  |
|  |  | Standard deviation | 0 | 1.3 | 2 | 2.2 |  |
|  |  | Median | 37.3 | 37.7 | 39.5 | 39.1 |  |
|  |  | Interquartile range | 37.3, 37.3 | 36.8, 38.7 | 39.5, 42.9 | 37.3, 39.5 |  |
|  |  | Negative value | 9 | 8 | 7 | 24 |  |
|  | ORF1a gene | N | 0 | 1 | 1 | 2 | n/a |
|  |  | Mean | n/a | 34 | 36.5 | 35.3 |  |
|  |  | Standard deviation | n/a | 0 | 0 | 1.8 |  |
|  |  | Median | n/a | 34 | 36.5 | 35.3 |  |
|  |  | Interquartile range | n/a | 34.0, 34.0 | 36.5, 36.5 | 34.0, 36.5 |  |
|  |  | Negative value | 10 | 9 | 9 | 28 |  |
|  | 18sRNA gene | N | 9 | 9 | 10 | 28 | 0.131 |
|  |  | Mean | 30 | 29.8 | 32.8 | 30.9 |  |
|  |  | Standard deviation | 2.9 | 3.9 | 4.6 | 4 |  |
|  |  | Median | 28.9 | 29.6 | 31 | 29.9 |  |
|  |  | Interquartile range | 28.5, 29.9 | 27.0, 31.2 | 29.9, 38.0 | 28.2, 31.4 |  |
|  |  | Negative value | 1 | 1 | 0 | 2 |  |
| Air sample (2nd) | E gene | N | 0 | 3 | 1 | 4 | n/a |
|  |  | Mean | n/a | 40.4 | 37.4 | 39.7 |  |
|  |  | Standard deviation | n/a | 2.8 | 0 | 2.8 |  |
|  |  | Median | n/a | 39.7 | 37.4 | 38.8 |  |
|  |  | Interquartile range | n/a | 38.0, 43.5 | 37.4, 37.4 | 37.7, 41.6 |  |
|  |  | Negative value | 10 | 7 | 9 | 26 |  |
|  | ORF1a gene | N | 0 | 1 | 0 | 1 | n/a |
|  |  | Mean | n/a | 37.2 | n/a | 37.2 |  |
|  |  | Standard deviation | n/a | 0 | n/a | 0 |  |
|  |  | Median | n/a | 37.2 | n/a | 37.2 |  |
|  |  | Interquartile range | n/a | 37.2, 37.2 | n/a | 37.2, 37.2 |  |
|  |  | Negative value | 10 | 9 | 10 | 29 |  |
|  | 18sRNA gene | N | 9 | 9 | 9 | 27 | 0.574 |
|  |  | Mean | 31.5 | 32.9 | 33.3 | 32.6 |  |
|  |  | Standard deviation | 3.9 | 4.9 | 4.5 | 4.4 |  |
|  |  | Median | 30.2 | 30.7 | 32.6 | 30.7 |  |
|  |  | Interquartile range | 29.0, 32.2 | 30.4, 32.8 | 29.8, 37.6 | 29.6, 35.9 |  |
|  |  | Negative value | 1 | 1 | 1 | 3 |  |
| Air sample (3rd) | E gene | N | 1 | 2 | 1 | 4 | 0.500 |
|  |  | Mean | 39.9 | 38.5 | 39.6 | 39.1 |  |
|  |  | Standard deviation | 0 | 1.4 | 0 | 1.1 |  |
|  |  | Median | 39.9 | 38.5 | 39.6 | 39.6 |  |
|  |  | Interquartile range | 39.9, 39.9 | 37.6, 39.5 | 39.6, 39.6 | 38.5, 39.7 |  |
|  |  | Negative value | 9 | 8 | 9 | 26 |  |
|  | ORF1a gene | N | 0 | 1 | 0 | 1 | n/a |
|  |  | Mean | n/a | 35.6 | n/a | 35.6 |  |
|  |  | Standard deviation | n/a | 0 | n/a | 0 |  |
|  |  | Median | n/a | 35.6 | n/a | 35.6 |  |
|  |  | Interquartile range | n/a | 35.6, 35.6 | n/a | 35.6, 35.6 |  |
|  |  | Negative value | 10 | 9 | 10 | 29 |  |
|  | 18sRNA gene | N | 10 | 10 | 10 | 30 | 0.583 |
|  |  | Mean | 31.8 | 32.1 | 32.5 | 32.1 |  |
|  |  | Standard deviation | 4.3 | 5.1 | 3.1 | 4.1 |  |
|  |  | Median | 30.2 | 30.1 | 31.3 | 30.3 |  |
|  |  | Interquartile range | 29.3, 34.3 | 28.6, 33.9 | 30.3, 35.9 | 29.4, 34.3 |  |
|  |  | Negative value | 0 | 0 | 0 | 0 |  |
| Surface sample (1st) | E gene | N | 3 | 2 | 1 | 6 | 0.100 |
|  |  | Mean | 36.9 | 39.2 | 34.4 | 37.3 |  |
|  |  | Standard deviation | 0.6 | 1.1 | 0 | 1.9 |  |
|  |  | Median | 37.3 | 39.2 | 34.4 | 37.3 |  |
|  |  | Interquartile range | 36.2, 37.3 | 38.4, 39.9 | 34.4, 34.4 | 36.2, 38.4 |  |
|  |  | Negative value | 7 | 8 | 9 | 24 |  |
|  | ORF1a gene | N | 4 | 2 | 2 | 8 | 0.210 |
|  |  | Mean | 37.8 | 39.4 | 37.6 | 38.1 |  |
|  |  | Standard deviation | 1.2 | 1.4 | 0.4 | 1.2 |  |
|  |  | Median | 38 | 39.4 | 37.6 | 38 |  |
|  |  | Interquartile range | 37.1, 38.5 | 38.4, 40.4 | 37.3, 37.8 | 37.6, 38.7 |  |
|  |  | Negative value | 6 | 8 | 8 | 22 |  |
|  | RNaseP gene | N | 9 | 9 | 10 | 28 | 0.658 |
|  |  | Mean | 33.7 | 33.1 | 33.2 | 33.3 |  |
|  |  | Standard deviation | 1 | 2 | 2 | 1.7 |  |
|  |  | Median | 33.8 | 33.3 | 33 | 33.3 |  |
|  |  | Interquartile range | 33.1, 34.2 | 32.0, 33.9 | 32.7, 34.2 | 32.6, 34.2 |  |
|  |  | Negative value | 1 | 1 | 0 | 2 |  |
| Surface sample (2nd) | E gene | N | 0 | 1 | 0 | 1 | n/a |
|  |  | Mean | n/a | 40 | n/a | 40 |  |
|  |  | Standard deviation | n/a | 0 | n/a | 0 |  |
|  |  | Median | n/a | 40 | n/a | 40 |  |
|  |  | Interquartile range | n/a | 40.0, 40.0 | n/a | 40.0, 40.0 |  |
|  |  | Negative value | 10 | 9 | 10 | 29 |  |
|  | ORF1a gene | N | 0 | 0 | 2 | 2 | n/a |
|  |  | Mean | n/a | n/a | 38.5 | 38.5 |  |
|  |  | Standard deviation | n/a | n/a | 0.9 | 0.9 |  |
|  |  | Median | n/a | n/a | 38.5 | 38.5 |  |
|  |  | Interquartile range | n/a | n/a | 37.8, 39.2 | 37.8, 39.2 |  |
|  |  | Negative value | 10 | 10 | 8 | 28 |  |
|  | RNaseP gene | N | 5 | 6 | 5 | 16 | 0.947 |
|  |  | Mean | 37.6 | 36.7 | 37.2 | 37.2 |  |
|  |  | Standard deviation | 0.6 | 2.4 | 3 | 2.1 |  |
|  |  | Median | 37.9 | 37.1 | 37.8 | 37.9 |  |
|  |  | Interquartile range | 37.8, 37.9 | 35.5, 38.7 | 36.7, 39.5 | 36.1, 38.7 |  |
|  |  | Negative value | 5 | 4 | 5 | 14 |  |
| Surface sample (3rd) | E gene | N | 2 | 1 | 1 | 4 | 0.500 |
|  |  | Mean | 38.4 | 34.8 | 39.4 | 37.8 |  |
|  |  | Standard deviation | 0.7 | 0 | 0 | 2.1 |  |
|  |  | Median | 38.4 | 34.8 | 39.4 | 38.4 |  |
|  |  | Interquartile range | 37.9, 39.0 | 34.8, 34.8 | 39.4, 39.4 | 36.4, 39.2 |  |
|  |  | Negative value | 8 | 9 | 9 | 26 |  |
|  | ORF1a gene | N | 0 | 1 | 0 | 1 | n/a |
|  |  | Mean | n/a | 35.4 | n/a | 35.4 |  |
|  |  | Standard deviation | n/a | 0 | n/a | 0 |  |
|  |  | Median | n/a | 35.4 | n/a | 35.4 |  |
|  |  | Interquartile range | n/a | 35.4, 35.4 | n/a | 35.4, 35.4 |  |
|  |  | Negative value | 10 | 9 | 10 | 29 |  |
|  | RNaseP gene | N | 3 | 4 | 3 | 10 | 0.791 |
|  |  | Mean | 37.4 | 36.5 | 37.6 | 37.1 |  |
|  |  | Standard deviation | 1.5 | 2 | 0.6 | 1.5 |  |
|  |  | Median | 36.6 | 37 | 37.9 | 37.1 |  |
|  |  | Interquartile range | 36.5, 39.2 | 35.2, 37.8 | 36.8, 38.0 | 36.6, 38.0 |  |
|  |  | Negative value | 7 | 6 | 7 | 20 |  |

## SUPPLEMENTARY FIGURES

**sFig. 1.**
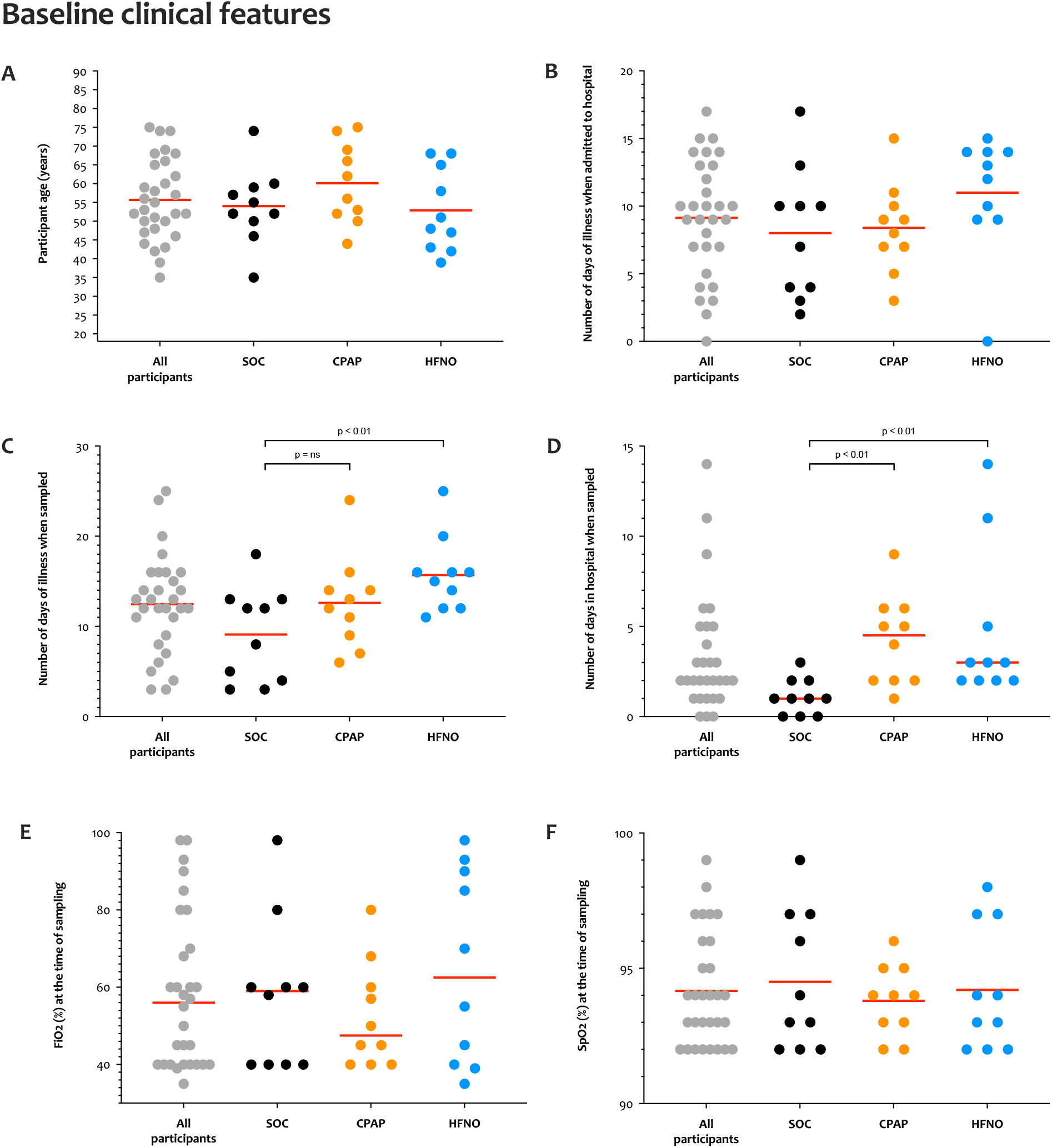
Baseline clinical characteristics of the study population. **(A) Study participant ages at time of enrolment.** The red bar denotes the mean. **(B) The number of days with COVID-19 symptoms at the time of hospital admission**. The red bar denotes the mean. **(C) The number of days with COVID-19 symptoms at the time of sampling**. The red bar denotes the mean. HFNO participants were sampled having been unwell for longer than SOC participants (mean SOC 9.1 days vs mean HFNO 15.7 days, p=0.01, two-tailed unpaired t-test). **(D) The number of days with in hospital at the time of sampling**. The red bar denotes the median. The median duration of hospital stay for SOC participants was one day, which was significantly lower than both CPAP and HFNO participants (median 4.5 and 3 days respectively, p<0.01 by two-tailed Mann-Whitney tests for CPAP and HFNO compared with SOC). **(E) % FiO**_**2**_ and **(F) SpO**_**2**_ at the time of sampling which did not differ between study groups (red bar denotes the median and mean respectively). SOC, supplemental oxygen care. CPAP, continuous positive airway pressure. HFNO, high-flow nasal oxygen. FiO_2_, fraction of inspired oxygen. SpO_2_, oxygen saturation.

**sFig. 2.**
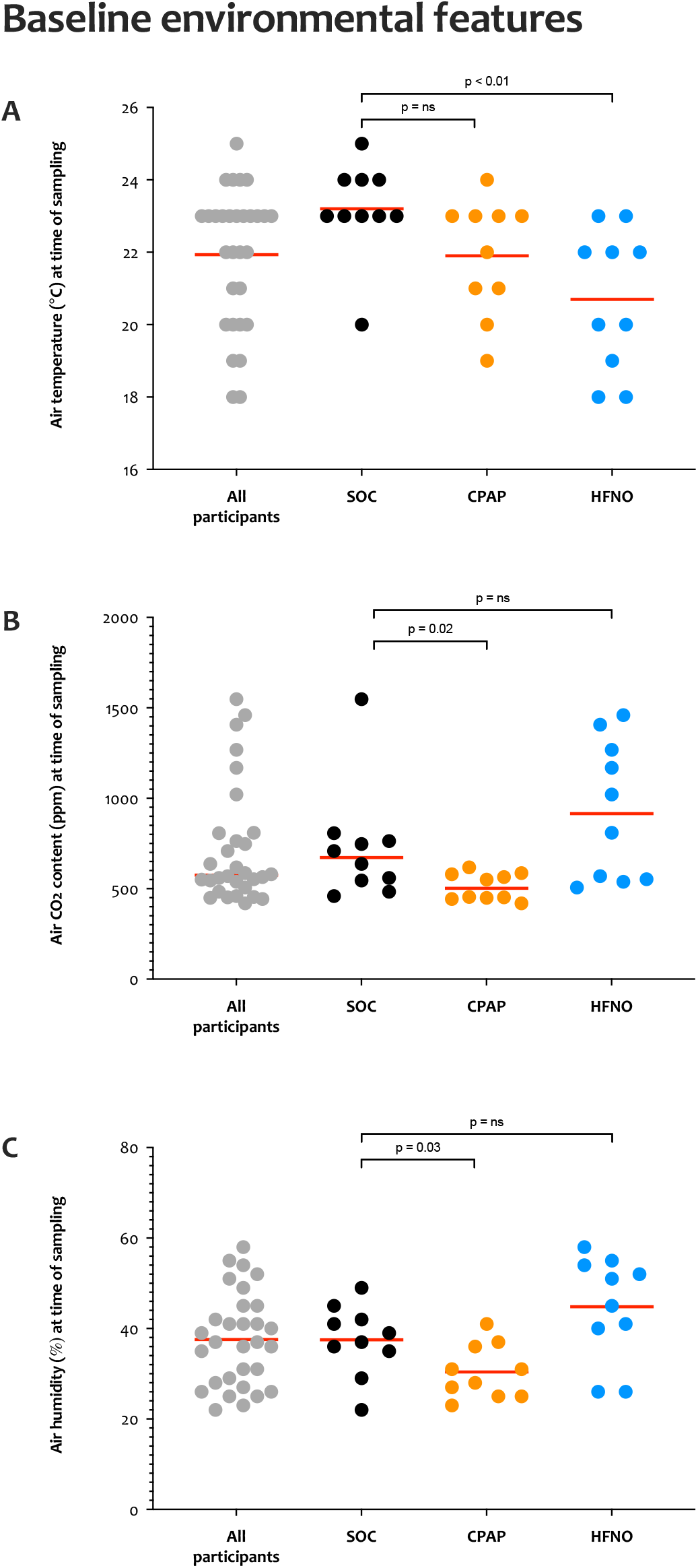
Baseline environmental characteristics of the clinical environments. **(A) Room air temperature**. The red bar denotes the mean. Air temperature from HFNO participants was lower than SOC participants (mean SOC 23.2°C vs mean HFNO 20.7°C, p<0.01, two-tailed unpaired t-test). **(B) Room air CO**_**2**_ **content**. The red bar denotes the median. The median CO_2_ content for SOC participants 673ppm, which was significantly higher than areas in use for CPAP (median 502ppm, p=0.02 by two-tailed Mann-Whitney tests). **(C) Room air humidity**. The red bar denotes the mean. The mean humidity of air around SOC participants was 37.6%, which was significantly higher than areas in use for CPAP (mean 30.4%, p=0.03 by two tailed unpaired t-test). SOC, supplemental oxygen care. CPAP, continuous positive airway pressure. HFNO, high-flow nasal oxygen.

**sFig. 3.**
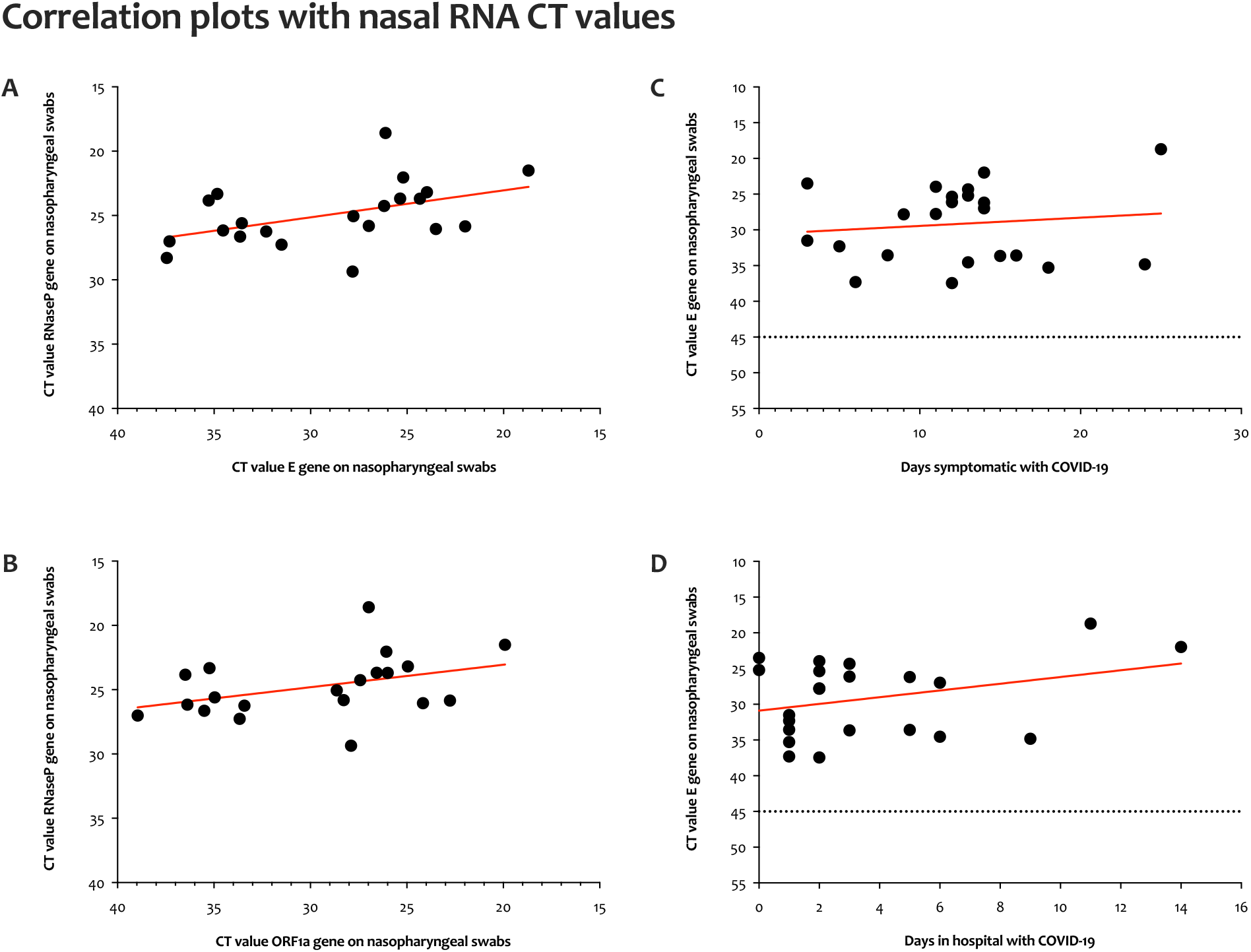
Nasopharyngeal human and viral RNA exploratory correlation plots. The red lines mark the linear regression analysis, dotted lines signify a Ct value of 45 that was used to define a negative result. **(A) Human RNaseP Ct values vs viral E gene Ct values and (B) Viral ORF1a gene Ct values. (C) Viral E gene Ct values vs days unwell with COVID-19 symptoms and (D) Days in hospital**. There was only a weak statistically significant correlation between human RNaseP Ct values and the viral E gene Ct value (r^2^=0.2, p=0.03, Pearson’s correlation), but not with ORF1a (r^2^=0.15, p=o.o9, Pearson’s correlation).

**sFig. 4.**
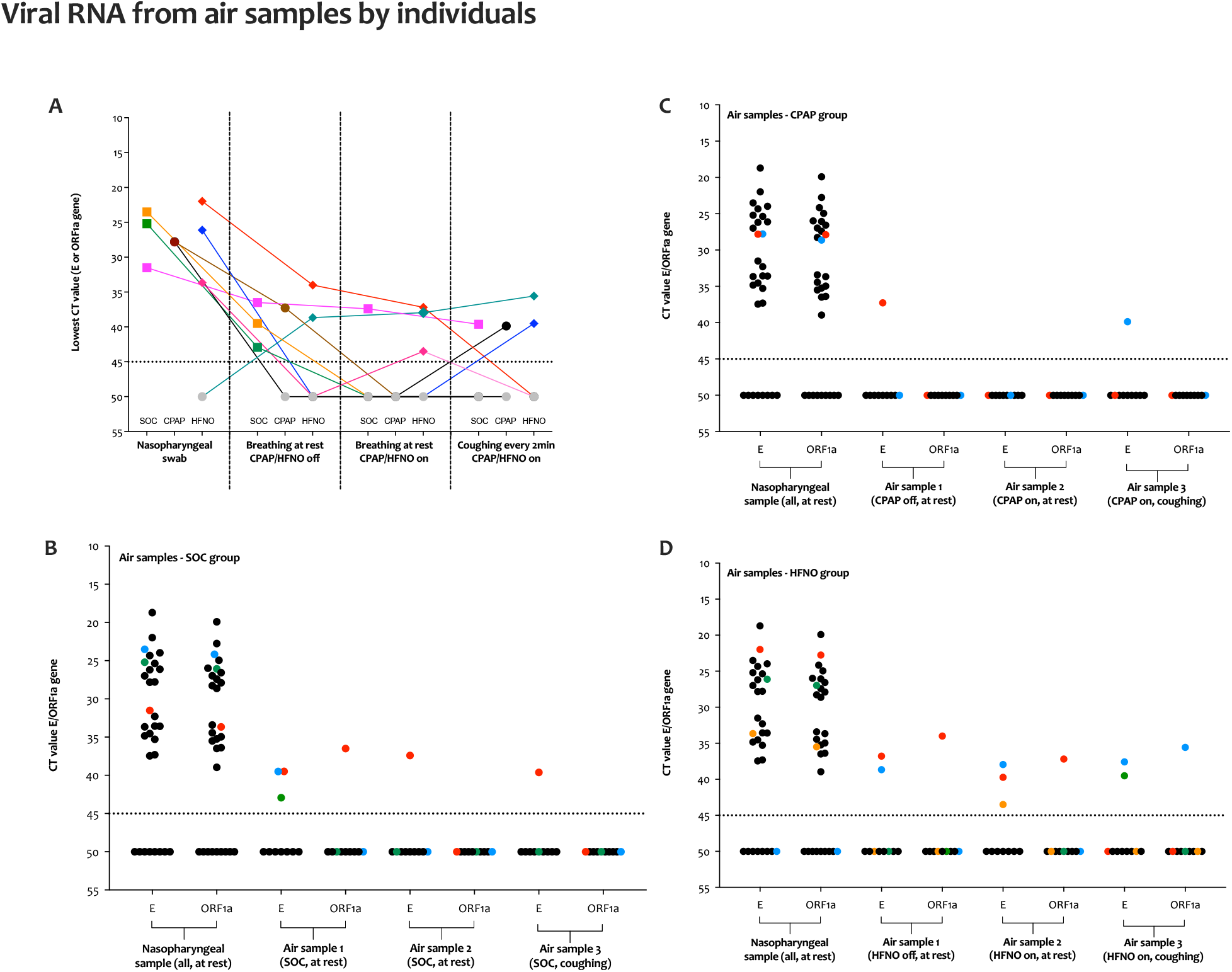
Viral RNA results from the nasopharynx and air samples linked as individual participants. The dotted lines signify a Ct value of 45 that was used to define a negative result. Ct values ≥ 45 were considered negative and were arbitrarily assigned a value of 50. **(A) All individuals linked from having had at least one positive or suspected-positive air sample**. The connected coloured dots are used to identify each participant with at least one positive Ct value for E/ORF1a genes (all study groups combined). **(B) SOC participants, (C) CPAP participants** and **(D) HFNO participants**. The coloured dots are used to identify each participant with at least one positive Ct value for E/ORF1a genes. SOC, supplemental oxygen care. CPAP, continuous positive airway pressure. HFNO, high-flow nasal oxygen.

**sFig. 5.**
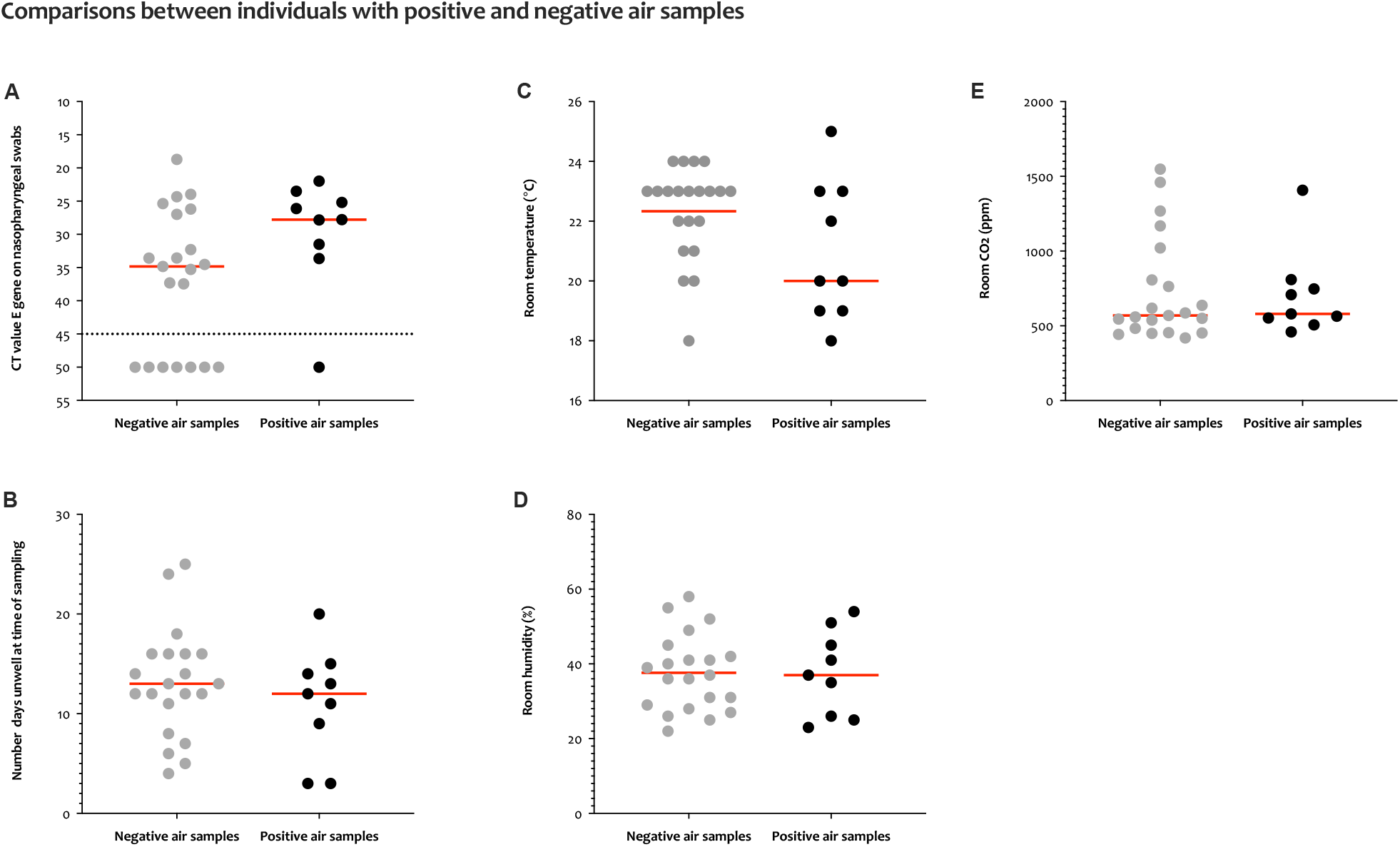
Sub-set comparative analysis of participants with positive and negative air samples. The red lines mark the mean or median Ct value as appropriate. Any single positive or suspected-positive air samples was used to identify the participants has having ‘positive air samples’ (n=9). The two sub-populations of participants were then compared for differences in the **(A) Ct values in the E gene from nasopharyngeal samples**. The dotted lines signify a Ct value of 45 that was used to define a negative result. **(B) Number of days unwell with COVID-19 symptoms** and environmental factors of **(C) Room air temperature, (D) Room air humidity** and **(E) Room air CO**_**2**_ **content**. Comparative testing (unpaired t-tests or Mann-Whitney tests, as appropriate) found no statistically significant differences between each of these sub-populations for any conditions shown above.

**sFig. 6.**
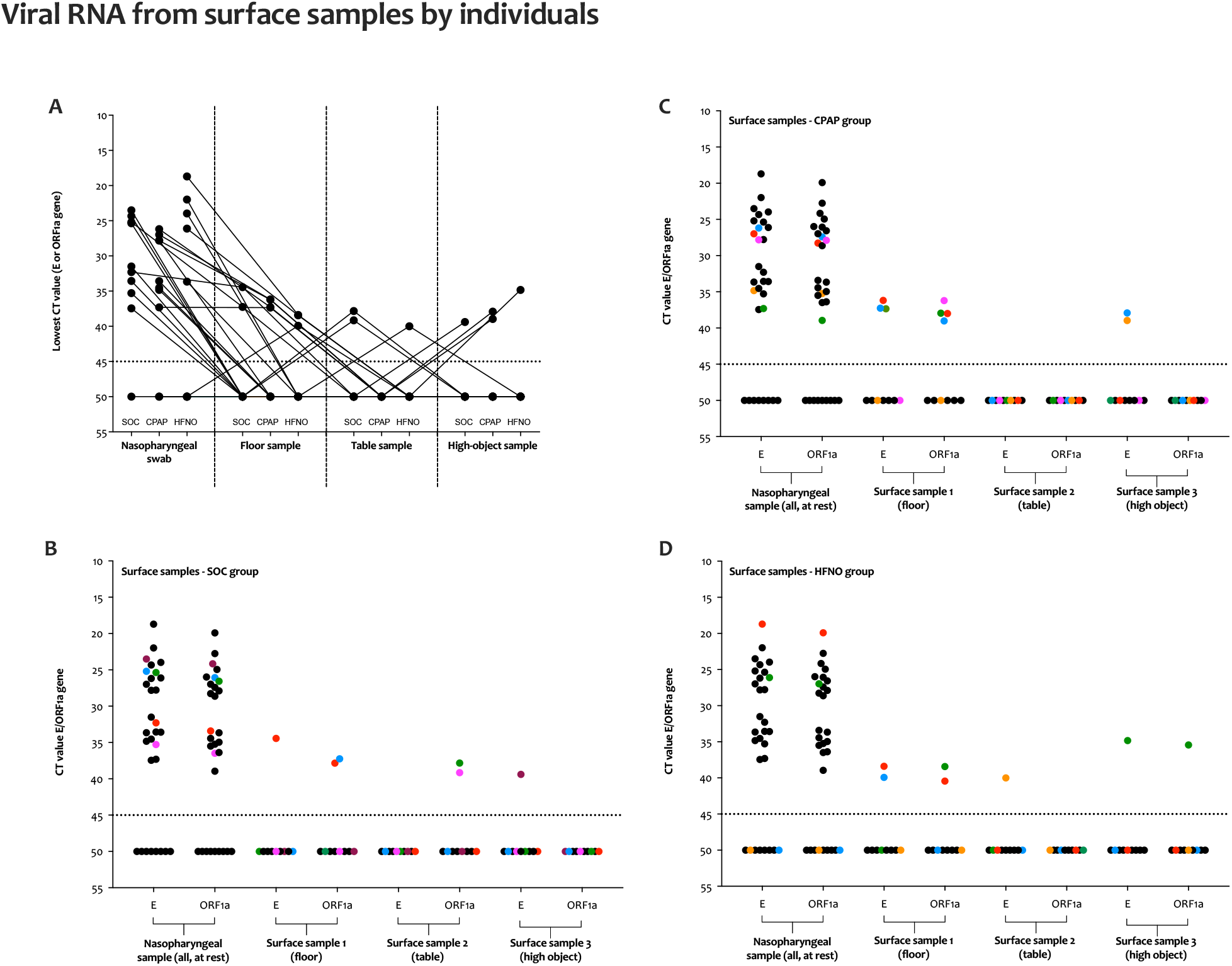
Viral RNA results from the nasopharynx and surface samples linked as individual participants. The dotted lines signify a Ct value of 45 that was used to define a negative result. Ct values ≥ 45 were considered negative and were arbitrarily assigned a value of 50. **(A) All individuals linked from having had at least one positive or suspected-positive surface sample**. The connected dots are used to identify each participant with at least one positive Ct value for E/ORF1a genes (all study groups combined). **(B) SOC participants, (C) CPAP participants** and **(D) HFNO participants**. The coloured dots are used to identify each participant with at least one positive Ct value for E/ORF1a genes. SOC, supplemental oxygen care. CPAP, continuous positive airway pressure. HFNO, high-flow nasal oxygen.

**sFig. 7.**
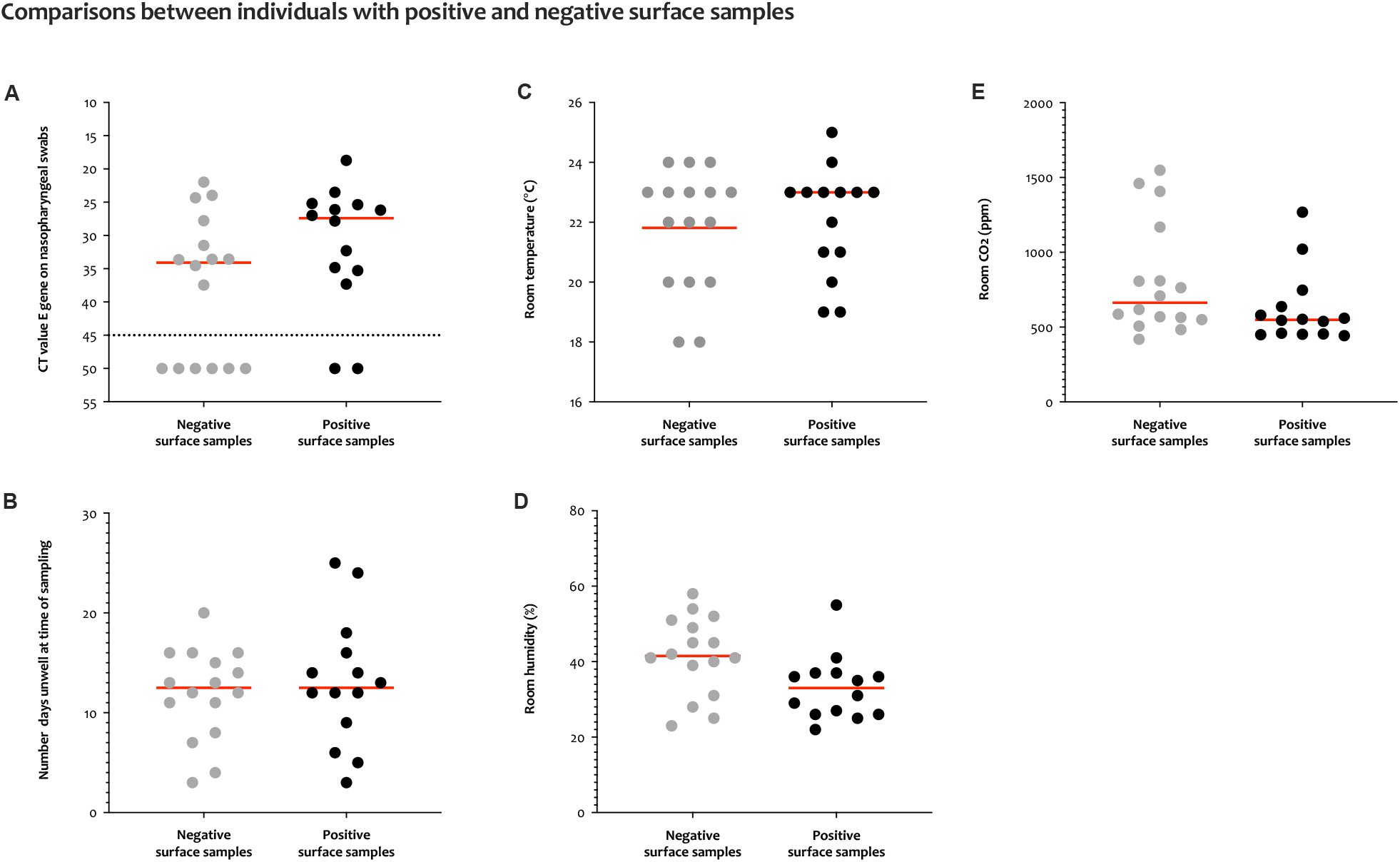
Sub-set comparative analysis of participants with positive and negative surface samples. The red lines mark the mean or median Ct value as appropriate. Any single positive or suspected-positive surface sample was used to identify the participants has having ‘positive surface samples’ (n=14). The two sub-populations of participants were then compared for differences in the **(A) Ct values in the E gene from nasopharyngeal samples**. The dotted lines signify a Ct value of 45 that was used to define a negative result. **(B) Number of days unwell with COVID-19 symptoms** and environmental factors of **(C) Room air temperature, (D) Room air humidity** and **(E) Room air CO**_**2**_ **content**. Comparative testing (unpaired t-tests or Mann-Whitney tests, as appropriate) found no statistically significant differences between each of these sub-populations for any conditions shown above, with the exception of room air humidity (p=0.02, two-tailed unpaired t-test). For participants in the positive/suspected-positive group for surface samples there was only a weak and non-significant correlation between room air humidity and the Ct value for viral E/ORF1a genes (r^2^=0.41, p=0.14, two-tailed Pearson’s correlation).

## NOTES

## Acknowledgements

The authors wish to especially thank the participants and their families who were willing to contribute towards this research, and whilst under very difficult personal circumstances. We also wish to thank the local R&D nurses and BHSF Occupational Health for supporting this work.

## Sponsor and funding

This report is independent research funded by the National Institute for Health Research. The views expressed in this publication are those of the author(s) and not necessarily those of the National Institute for Health Research or the Department of Health and Social Care. The University of Oxford is the Sponsor for the ISARIC WHO CCP-UK protocol.

## Ethical approvals and study registrations

Ethical approval and amendments for ISARIC was granted by South Central Oxford C Research Ethics Committee (reference 13/SC/0149). ISARIC WHO CCP-UK study registration ISRCTN 66726260. Written informed consent was obtained from all volunteers prior to study procedures. Study procedures were conducted at Birmingham Heartlands Hospital, Good Hope Hospital and the Queen Elizabeth Hospital Birmingham (part of University Hospitals Birmingham NHS Foundation Trust). Procedures were performed in accordance with ICH Good Clinical Practice (GCP) and local standard operating procedures (SOPs). The study was monitored by the Research Governance team at University Hospitals Birmingham NHS Foundation Trust.

## Author contributions

RLW was the lead physician, analysed the data and wrote the manuscript. DMcA, GP and CAG instigated and designed the study. Sample collection and processing was performed by RLW, EW, IN, JZ and CAG. Laboratory analyses were performed by JZ and WB. Statistical analyses were performed by CJ, RL, RLW and CAG. All authors have reviewed and had input into the manuscript prior to submission.

## Competing interests

All authors have completed the ICMJE uniform disclosure form at www.icmje.org/coi_disclosure.pdf and have no competing interests to declare.

## Notes

### Competing Interest Statement

The authors have declared no competing interest.

### Author Declarations

Ethical approval Oxford C Research Ethics Committee (13/SC/0149).

